# Psychosocial predictors of COVID-19 vaccine uptake among pregnant women: a cross-sectional study in Greece

**DOI:** 10.1101/2022.04.06.22273526

**Authors:** Petros Galanis, Irene Vraka, Aglaia Katsiroumpa, Olympia Konstantakopoulou, Olga Siskou, Eleftheria Zogaki, Daphne Kaitelidou

**Author notes:** **Corresponding author:** Petros Galanis, Assistant Professor, Clinical Epidemiology Laboratory, Faculty of Nursing, National and Kapodistrian University of Athens, 123 Papadiamantopoulou street, GR-11527, Athens, Greece.

## Abstract

**Background:** Unvaccinated pregnant women with symptomatic COVID-19 have been found to have a higher risk of iatrogenic preterm births, intensive care unit admission, and invasive ventilation.

**Objective:** To estimate the vaccination rate of pregnant women against the COVID-19 and to evaluate psychosocial factors associated with vaccine uptake among them.

**Methods:** We conducted an anonymous cross-sectional study with a convenience sample in Greece from December 2021 to March 2022. We measured socio-demographic data of pregnant women, COVID-19-related vaccination status, worry about the side effects of COVID-19 vaccines, trust in COVID-19 vaccines, and COVID-19-related stress.

**Results:** The study population included 812 pregnant women with a mean age of 31.6 years. Among the pregnant women, 58.6% had received a COVID-19 vaccine. The most important reasons that pregnant women were not vaccinated were doubts about the safety and effectiveness of COVID-19 vaccines (31.4%), fear that COVID-19 vaccines could be harmful to fetus (29.4%), and fear of adverse side effects of COVID-19 vaccines (29.4%). Increased danger and contamination fears, increased fears about economic consequences, and higher levels of trust in COVID-19 vaccines were related with COVID-19 vaccine uptake. On the other hand, increased compulsive checking and reassurance seeking and increased worry about the adverse side effects of COVID-19 vaccines reduced the likelihood of pregnant women being vaccinated against the COVID-19.

**Conclusions:** An understanding of the psychosocial factors associated with COVID-19 vaccine uptake in pregnant women is paramount to persuade women to get vaccinated against the COVID-19. There is a need for targeted educational campaigns to increase knowledge about COVID-19 vaccines and reduce COVID-19 vaccine hesitancy in pregnancy.

## Introduction

Unvaccinated pregnant women with symptomatic COVID-19 are at increased risk for iatrogenic preterm births, intensive care unit admission, and invasive ventilation (Khalil et al., 2020; Lokken et al., 2021; Vousden et al., 2021). Also, risk of death for unvaccinated women with symptomatic COVID-19 during pregnancy is higher than non pregnant women with symptomatic COVID-19 (Zambrano et al., 2020). Moreover, unvaccinated pregnant women have been found to have a higher risk of hospital admission for COVID-19 than vaccinated pregnant women (Iacobucci, 2021).

Pregnant women were excluded from early randomized controlled trials, leading to the lack of safety and efficacy data (Pogue et al., 2020; Polack et al., 2020). However, the most recent data are rather encouraging for vaccinated pregnant women. In particular, COVID-19 vaccine research finds that (1) abortion rate, adverse pregnancy, and adverse neonatal outcomes are similar in vaccinated and non-vaccinated pregnant women, (2) anti-SARS-CoV-2 immunoglobulins provide immunity to the newborns, and (3) COVID-19 vaccines do not cause vaccine-related adverse events (Falsaperla et al., 2021; Fu et al., 2022; Garg et al., 2021; Mendoza et al., 2020; Rawal et al., 2022). Thus, several organizations worldwide, now recommend COVID-19 vaccination for pregnant women and women who are trying to become pregnant, or who might become pregnant in the future to prevent severe maternal morbidity and adverse birth outcomes (American College of Obstetricians and Gynecologist, 2021; Centers for Disease Control and Prevention, 2021).

Until now, only a handful of studies investigated the determinants of COVID-19 vaccine uptake among pregnant women (Blakeway et al., 2022; Hosokawa et al., 2022; Razzaghi et al., 2021; Siegel et al., 2021; UK Health Security Agency, 2021). Moreover, only one study searched for psychosocial predictors of COVID-19 vaccine uptake among pregnant women, e.g. trust in COVID-19 vaccines, fear of COVID-19, worry about the adverse side effects of the COVID-19 vaccines, etc. (Siegel et al., 2021). So far, research has focused on the impact of the socio-demographic characteristics on the decision of pregnant women to be vaccinated against the COVID-19. Thus, the aim of our study was to estimate the rate of vaccinated pregnant women against the COVID-19 and to study psychosocial predictors of COVID-19 vaccine uptake.

## Methods

### Study design and participants

We conducted a cross-sectional study with a convenience sample in Greece. Recruitment of pregnant women began about one year after the Greek government has offered a free COVID-19 vaccine to all adults. In particular, we collected data from December 2021 to March 2022. We used several approaches to collect our data. Firstly, we used Google forms to create an anonymous version of the study questionnaire and then we disseminated it through social media, e.g. Facebook. Moreover, we sent the questionnaire via e-mail to all our contacts targeting in pregnant women. At the same time, we asked pregnant women who completed the study questionnaire to invite to our study any other pregnant women they might have known. Thus, a snowball sampling method was applied. Women were considered eligible for the study if they were pregnant aged 18 or older and indicated that they could complete the study questionnaire in Greek. Data were anonymous and confidential and participation in the survey was voluntary. We informed the pregnant women that they could cease to participate at any stage of the study should they wish to do so.

Considering a low effect size (odds ratio=1.4), the precision level as 5% (alpha level), and the power as 95%, a minimum sample size of 564 pregnant women was required. Nevertheless, we strove to recruit a larger number of pregnant women to further decrease random error.

The Ethics Committee of Department of Nursing, National and Kapodistrian University of Athens approved the study protocol (reference number; 370, 02-09-2021).

### Questionnaires

Firstly, we collected the following socio-demographic data of pregnant women: age (continuous), marital status (singles, married, in a couple relationship without marriage, or divorced), educational level (elementary school, high school, or university degree), financial status (very poor, poor, average, good, or very good), physical health (very poor, poor, average, good, or very good), children aged 18 or younger (no or yes), chronic disease (no or yes), previous COVID-19 diagnosis (no or yes), and family members/friends with previous COVID-19 diagnosis (no or yes).

Also, we used four questions to measure pregnant women’ vaccination status and COVID-19-related vaccination status. In particular, we asked the pregnant women (1) if they have received the vaccination against the seasonal influenza (no or yes), (2) if they have received the vaccination against the COVID-19 (no or yes), and (3) if they have received the vaccination against the COVID-19 before or during pregnancy (no or yes). Moreover, we asked unvaccinated pregnant women to state the main reason for this denial (doubts about the safety and effectiveness of COVID-19 vaccines, fear of adverse side effects of COVID-19 vaccines, low self-perceived threat regarding COVID-19, previous COVID-19 diagnosis, fear due to a chronic disease, family doctor’ recommendation due to physical health of pregnant women, religious issues, or fear that the COVID-19 vaccination could be harmful to the fetus).

We measured pregnant women’s worry about the side effects of COVID-19 vaccines with a single item: “I am worried about the side effects that COVID-19 vaccines can have”. Also, we measured pregnant women’s trust in COVID-19 vaccines with a single item: “I trust the COVID-19 vaccines”. Answers in both cases were indicated on a scale ranging from 0 (totally disagree) to 10 (totally agree).

We used the 36-item COVID Stress Scales (CSS) to measure COVID-19-related stress (Taylor et al., 2020). In particular, we used the Greek version of the CSS (Galanis, Vraka, Katsiroumpa, et al., 2022). The questionnaire consists of five factors: (1) Danger and contamination fears, (2) fears about economic consequences, (3) xenophobia, (4) compulsive checking and reassurance seeking, and (5) traumatic stress symptoms about COVID-19. Responses for the first three factors are indicated on a 5-point Likert scale ranging from 0 (not at all) to 4 (extremely), and for the last two factors are indicated on a 5-point Likert scale ranging from 0 (never) to 4 (almost always). For each factor, a stress score is calculated by averaging the answers to all items of the factor. Thus, score for each factor ranges from 0 to 4 with higher scores reflect greater stress. In our study, Cronbach’s alpha was 0.93, 0.94, 0.95, 0.88, and 0.93 for the five factors respectively.

### Statistical analysis

We use numbers and percentages to present categorical variables and mean and standard deviation to present continuous variables. Socio-demographic data of pregnant women, COVID-19-related stress, worry about the adverse side effects of COVID-19 vaccines, and trust in COVID-19 vaccines were the independent variables in our study. The outcome variable was COVID-19 vaccination uptake among pregnant women, measured through “yes/no” answers. Thus, we used logistic regression analysis to identify the predictors of COVID-19 vaccine uptake. We performed univariate and multivariate logistic regression analysis, calculating unadjusted and adjusted odds ratios respectively. Also, we calculated 95% confidence intervals (CI), and two-sided p-values. A p-value<0.05 considering as statistically significant. Statistical analysis was performed with the Statistical Package for Social Sciences software (IBM Corp. Released 2012. IBM SPSS Statistics for Windows, Version 21.0. Armonk, NY: IBM Corp.).

## Results

### Socio-demographic characteristics

The study population included 812 pregnant women. Mean age of the pregnant women was 31.6 years, while 35.3% were expecting their first child. The majority of the participants were married or in a couple relationship without marriage (89.7%). Among pregnant women, 48.3% defined their financial status as good/very good. Almost 15% of the pregnant women suffered from a chronic disease and 90.5% defined their health status as good/very good. Twenty-four point one percent of pregnant women and 79.3% of their family members/friends were diagnosed with COVID-19. Detailed socio-demographic characteristics of the pregnant women are shown in Table 1.

**Table 1.** Socio-demographic characteristics of the pregnant women.

| **Variable** | **N** | **%** |
| --- | --- | --- |
| Marital status |  |  |
| Singles | 84 | 10.3 |
| Married or in a couple relationship without marriage | 728 | 89.7 |
| Divorced | 0 | 0 |
| Age (years)<sup>a</sup> | 31.6 | 4.6 |
| Children <18 years old |  |  |
| No | 525 | 64.7 |
| Yes | 287 | 35.3 |
| Educational level |  |  |
| Elementary school | 7 | 0.9 |
| High school | 210 | 25.9 |
| University degree | 595 | 73.3 |
| Self-perceived financial status |  |  |
| Very poor | 7 | 0.9 |
| Poor | 28 | 3.4 |
| Moderate | 385 | 47.4 |
| Good | 336 | 41.4 |
| Very good | 56 | 6.9 |
| Self-perceived health status |  |  |
| Very poor | 0 | 0 |
| Poor | 0 | 0 |
| Moderate | 77 | 9.5 |
| Good | 462 | 56.9 |
| Very good | 273 | 33.6 |
| Chronic disease |  |  |
| No | 693 | 85.3 |
| Yes | 119 | 14.7 |
| Previous COVID-19 diagnosis |  |  |
| No | 616 | 75.9 |
| Yes | 196 | 24.1 |
| Family members/friends with previous COVID-19 diagnosis |  |  |
| No | 168 | 20.7 |
| Yes | 644 | 79.3 |

<sup>a</sup> mean, standard deviation

### COVID-19-related vaccination status

Among the pregnant women in our study, 58.6% had received a COVID-19 vaccine. Among the vaccinated women, 45.6% had received a COVID-19 vaccine during pregnancy. The most important reasons that pregnant women were not vaccinated were doubts about the safety and effectiveness of COVID-19 vaccines (31.4%), fear that COVID-19 vaccines could be harmful to my fetus (29.4%), and fear of adverse side effects of COVID-19 vaccines (29.4%). In our sample, 24.1% of the pregnant women had received a flu vaccine during 2021. COVID-19-related vaccination status of the pregnant women is presented in Table 2.

**Table 2.** COVID-19-related vaccination status in the pregnant women.

| **Variable** | **N** | **%** |
| --- | --- | --- |
| COVID-19 vaccination uptake |  |  |
| No | 336 | 41.4 |
| Yes | 476 | 58.6 |
| COVID-19 vaccination uptake before pregnancy |  |  |
| No | 217 | 45.6 |
| Yes | 259 | 54.4 |
| Seasonal influenza vaccination in 2021 |  |  |
| No | 616 | 75.9 |
| Yes | 196 | 24.1 |
| Reasons for decline of pregnant women to receive a COVID-19 vaccine |  |  |
| I have doubts about the safety and effectiveness of COVID-19 vaccines | 112 | 31.4 |
| I am afraid of adverse side effects of COVID-19 vaccines | 105 | 29.4 |
| I believe that even I get infected with COVID-19, nothing bad will happen to me | 14 | 3.9 |
| I have already been diagnosed with COVID-19 and the vaccine will not be beneficial for me | 14 | 3.9 |
| I am afraid because I suffer from a chronic disease | 7 | 2.0 |
| I am afraid that COVID-19 vaccines could be harmful to my fetus | 105 | 29.4 |

### COVID-19-related stress

The highest levels of COVID-19-related stress were due to danger and contamination fears and then to xenophobia, compulsive checking and reassurance seeking, fears about economic consequences, and traumatic stress symptoms about COVID-19 was low. The level of danger and contamination fears was moderate for both vaccinated and unvaccinated pregnant women, while the level of (1) fears about economic consequences, (2) xenophobia, (3) compulsive checking and reassurance seeking, and (4) traumatic stress symptoms about COVID-19 was low. The mean level of danger and contamination fears, fears about economic consequences, xenophobia, compulsive checking and reassurance seeking, and traumatic stress symptoms about COVID-19 among the vaccinated pregnant women is higher than that of the unvaccinated pregnant women. COVID-19-related stress of pregnant women according to COVID-19 vaccination status is shown in Table 3.

**Table 3.** COVID-19-related stress of pregnant women according to COVID-19 vaccination status.

| **Scale** | **Mean** | **Standard deviation** |
| --- | --- | --- |
| Danger and contamination fears |  |  |
| Unvaccinated | 2.01 | 1.06 |
| Vaccinated | 2.34 | 1.02 |
| Fears about economic consequences |  |  |
| Unvaccinated | 0.51 | 0.82 |
| Vaccinated | 0.65 | 1.05 |
| Xenophobia |  |  |
| Unvaccinated | 0.97 | 1.24 |
| Vaccinated | 1.04 | 1.27 |
| Compulsive checking and reassurance seeking |  |  |
| Unvaccinated | 0.87 | 0.84 |
| Vaccinated | 0.92 | 0.97 |
| Traumatic stress symptoms about COVID-19 |  |  |
| Unvaccinated | 0.40 | 0.74 |
| Vaccinated | 0.45 | 0.74 |

### Predictors of COVID-19 vaccines uptake

Multivariate logistic regression analysis identified that five psychosocial factors and seven socio-demographic characteristics affect pregnant women decision to receive a COVID-19 vaccine. The multivariate logistic regression analysis for COVID-19 vaccine uptake among the pregnant women revealed that the independent variables explained 55.6% of the variance in this outcome. Predictors of COVID-19 vaccines uptake are shown in Table 4.

**Table 4.** Univariate and multivariate logistic regression analysis with COVID-19 vaccine uptake among the pregnant women as the dependent variable (reference: COVID-19 vaccine denial).

| Variable | Unadjusted OR (95% CI) | P-value | Adjusted OR (95% CI)<sup>a</sup> | P-value |
| --- | --- | --- | --- | --- |
| Marital status (singles vs. married) | 1.45 (0.91 – 2.36) | 0.12 | 2.43 (1.09 – 5.43) | **0.03** |
| Age (years) | 1.07 (1.04 – 1.10) | <0.001 | 1.09 (1.04 – 1.15) | **0.001** |
| Children aged <18 years old (yes vs. no) | 1.36 (1.01 – 1.83) | 0.04 | 1.84 (1.19 – 2.83) | **0.006** |
| Educational level (University degree vs. high school) | 2.75 (1.99 – 3.79) | <0.001 | 1.08 (0.65 – 1.77) | 0.77 |
| Self-perceived financial status (very good/good vs. moderate/poor/very poor) | 1.18 (0.89 – 1.56) | 0.24 | 0.91 (0.59 – 1.39) | 0.67 |
| Self-perceived health status (very poor/poor/moderate vs. good/ very good) | 1.26 (0.78 – 2.05) | 0.35 | 3.17 (1.49 – 6.72) | **0.003** |
| Chronic disease (yes vs. no) | 1.35 (0.90 – 2.03) | 0.15 | 0.95 (0.52 – 1.73) | 0.85 |
| Previous COVID-19 diagnosis (no vs. yes) | 1.31 (0.95 – 1.81) | 0.10 | 1.69 (1.04 – 2.78) | **0.04** |
| Family members/friends with previous COVID-19 diagnosis (yes vs. no) | 2.39 (1.69 – 3.38) | <0.001 | 1.82 (1.09 – 3.05) | **0.02** |
| Seasonal influenza vaccination in 2021 (yes vs. no) | 8.72 (5.40 – 14.09) | <0.001 | 17.89 (8.51 – 37.64) | **<0.001** |
| Danger and contamination fears | 1.35 (1.18 – 1.54) | <0.001 | 1.38 (1.07 – 1.78) | **0.013** |
| Fears about economic consequences | 1.16 (0.99 – 1.35) | 0.05 | 1.48 (1.09 – 2.01) | **0.01** |
| Xenophobia | 1.04 (0.93 – 1.17) | 0.47 | 1.07 (0.85 – 1.35) | 0.54 |
| Compulsive checking and reassurance seeking | 1.07 (0.91 – 1.24) | 0.41 | 0.46 (0.35 – 0.62) | **<0.001** |
| Traumatic stress symptoms about COVID-19 | 1.09 (0.89 – 1.32) | 0.38 | 1.19 (0.86 – 1.66) | 0.28 |
| Worry about the side effects of COVID-19 vaccines | 0.84 (0.81 – 0.88) | <0.001 | 0.73 (0.67 – 0.78) | **<0.001** |
| Trust in COVID-19 vaccines | 1.36 (1.29 – 1.44) | <0.001 | 1.53 (1.41 – 1.66) | **<0.001** |

An odds ratio <1 indicates a negative association, while an odds ratio >1 indicates a positive association. Bold values indicate statistically significant relations in multivariate model.
CI: confidence interval; OR: odds ratio
<sup>a</sup> R<sup>2</sup> for the final multivariate model was 55.6%

Regarding psychosocial factors, increased danger and contamination fears, increased fears about economic consequences, and higher levels of trust in COVID-19 vaccines were related with pregnant women’s COVID-19 vaccine uptake. On the other hand, increased compulsive checking and reassurance seeking and increased worry about the adverse side effects of COVID-19 vaccines reduced the likelihood of pregnant women being vaccinated against the COVID-19.

Regarding socio-demographic characteristics, singles women, women that already had at least one child aged <18 years old, women that defined their health status as very poor/poor/moderate, women without a previous COVID-19 diagnosis, women with family members/friends with previous COVID-19 diagnosis, and women who were vaccinated against the seasonal influenza had a greater probability to be vaccinated against the COVID-19. Moreover, increased age was significantly associated with higher COVID-19 vaccine uptake among pregnant women.

## Discussion

To the best of our knowledge this is the first study that investigates psychosocial predictors of COVID-19 vaccine uptake among pregnant women with a valid questionnaire. Siegel et al. 2021 studied several psychosocial predictors but without using a valid questionnaire. In a sample of pregnant women in Greece, we found that 58.6% of them were vaccinated against the COVID-19 during pregnancy. A recent meta-analysis found that the overall proportion of vaccinated pregnant women against the COVID-19 is 27.5% (Galanis, Vraka, Siskou, Konstantakopoulou, Katsiroumpa, & Kaitelidou, 2022b). Also, it is interesting to highlight the fact that the vaccination rate is much higher in Israel (43.3%) than in the USA (27.3%) and other countries (12.8%). The higher vaccination rate in our study may be due to the fact that the studies in the meta-analysis were conducted from December 2020 to September 2021, while our study was conducted from December 2021 to March 2022. Evidence on the safety and efficacy of the COVID-19 vaccines is constantly increasing which may also increase the confidence of pregnant women in vaccines.

We found that the main reasons that pregnant women were not vaccinated were doubts about the safety and effectiveness of COVID-19 vaccines, fear that COVID-19 vaccines could be harmful to my fetus, and fear of adverse side effects. This finding is confirmed by the literature, since several studies have revealed women’s fear that COVID-19 vaccines can cause problems in fertility, pregnancy, and breastfeeding (Nguyen et al., 2021; Schrading et al., 2021; Xu et al., 2021). Moreover, pregnant women are more worried about lack of safety data in pregnancy, fetal effects, vaccine adverse side effects, and rushed development of vaccines (Siegel et al., 2021; Skjefte et al., 2021). Moreover, worry in the general population about safety, efficacy, and side-effects of COVID-19 vaccines is related with hesitancy in COVID-19 vaccines uptake (Freeman et al., 2020).

According to our multivariate analysis, several psychosocial factors affect pregnant women decision to receive a COVID-19 vaccine. In particular, increased danger, contamination fears, and fears about economic consequences were related with COVID-19 vaccine uptake among pregnant women. Siegel et al. (2021) confirm our finding, since they found that vaccinated pregnant women are more afraid of the COVID-19 during pregnancy than unvaccinated pregnant women. Moreover, vaccinated pregnant women believe that if they are infected they are at risk for getting very sick (Siegel et al., 2021). Increased fear of pregnant women in our study may also be due to the fact that they have family members/friends with previous COVID-19 diagnosis. Siegel et al. (2021) confirm this result since vaccinated pregnant women are more likely to know hospitalized COVID-19 patients. In general, fear of COVID-19 is associated with good prevention practices in both pregnant women and the general population (Ahorsu et al., 2022; Masjoudi et al., 2022; W/Mariam et al., 2021; Wong et al., 2020; Yildirim et al., 2021). Probably, individuals who fear to acquire and spread the COVID-19 could better adopt a protective health behavior against the COVID-19 and comply with preventive measures recommendations. Therefore, fear appears to promote preventive measures during the COVID-19 pandemic, e.g. frequent hand-washing, physical distancing, face masks, and vaccination.

On the other hand, we found that high levels of COVID-19-related stress and more specifically the stress caused by the compulsive checking and reassurance seeking reduce the COVID-19 vaccine uptake among the pregnant women. Pregnant women experience moderate to high levels of stress during the COVID-19 pandemic (Ayaz et al., 2020; Mappa et al., 2020; Stepowicz et al., 2020). Particular attention should be focused on pregnant women since COVID-19-related stress is associated with depressive symptoms (King et al., 2021). Higher depressive symptoms in pregnancy may have negative effects on infants’ health, e.g. disruptions of pro- and anti-inflammatory markers, infant stress responses, shortened breastfeeding period, and poorer bonding between mothers and their infants (Edvinsson et al., 2017; Figueiredo et al., 2014; Gustafsson et al., 2018; Osborne et al., 2018; Rossen et al., 2016). Moreover, there is a negative association between perceived stress and self-care behaviors among pregnant women (Masjoudi et al., 2022). Negative self-care behaviors among pregnant women with high levels of COVID-19-related stress could explain the low vaccination rate among those women since self-care behaviors are purposeful actions that individuals take on to improve their health (Masjoudi et al., 2020; Stearns et al., 2000).

Also, we found that higher levels of trust in COVID-19 vaccines were associated with higher COVID-19 vaccine uptake among pregnant women. Literature confirms this finding, since Siegel et al. (2021) found that trust in COVID-19 vaccines effectiveness for women and newborns is associated with increased vaccine uptake. Also, they recognized that unvaccinated pregnant women are less likely to trust vaccine developers. Moreover, general distrust is related with hesitancy in COVID-19 vaccines uptake in the general population (Freeman et al., 2020). It is noteworthy that the mistrust in the COVID-19 vaccines is higher among individuals from ethnic minorities, both in the general population and among pregnant women (Green et al., 2021; Paul et al., 2021; Taubman – BeneAri et al., 2022). Another important issue is that several studies found a positive relation between trust in COVID-19 vaccines and parents’ willingness to vaccinate their children against the COVID-19 (Galanis, Vraka, Siskou, Konstantakopoulou, Katsiroumpa, & Kaitelidou, 2022a; Skjefte et al., 2021; Yılmaz & Sahin, 2021; Zhang et al., 2020).

Our study identified several socio-demographic predictors of COVID-19 vaccination uptake among pregnant women. Among others, we found that pregnant women who received a flu vaccine had also a greater probability to be vaccinated against the COVID-19. Systematic reviews have already shown that previous seasonal influenza vaccination history is a strong predictive factor for individuals to accept a COVID-19 vaccine, both for themselves and their children (Galanis, Vraka, et al., 2021; Galanis, Vraka, Siskou, Konstantakopoulou, Katsiroumpa, & Kaitelidou, 2022a; Li et al., 2021; Luo et al., 2021; Shamshirsaz et al., 2021). Moreover, recent studies expand this evidence by confirming that COVID-19 vaccination uptake is more common among individuals that are already vaccinated against the seasonal influenza (Doran et al., 2022; Galanis, Moisoglou, et al., 2021; Galanis, Vraka, Siskou, Konstantakopoulou, Katsiroumpa, Moisoglou, et al., 2022; Gilboa et al., 2021; Shariff et al., 2022). Influenza vaccination during the COVID-19 pandemic is crucial since may have a protective role against the COVID-19 pandemic by reducing SARS-CoV-2 infection rate, hospitalization, severity of COVID-19, admission to intensive care unit and mortality (Wang et al., 2021; Zeynali Bujani et al., 2021).

Older age of pregnant women is another predictor of COVID-19 vaccination uptake. Our findings confirm the evidence that older age is associated with increased likelihood of acceptance and uptake of a COVID-19 vaccine among pregnant women (Razzaghi et al., 2021; Skjefte et al., 2021; Stuckelberger et al., 2021; UK Health Security Agency, 2021). Probably, older pregnant women experience more fear of COVID-19 since the evidence shows that pregnancy at advanced maternal age is a strong predictor for adverse outcomes, e.g. neonatal intensive care unit admission, low birthweight babies, worse Apgar scores, spontaneous miscarriage, cesarean deliveries, and pre-eclampsia (Glick et al., 2021; Pinheiro et al., 2019). Moreover, it is well known that older age increases adverse outcomes in COVID-19 patients, such as hospitalization, intensive care unit admission and mortality (Mehraeen et al., 2020; Sepandi et al., 2020; Yanez et al., 2020). This fear that pregnant women experience against the COVID-19 is confirmed by two more finding in our study. In particular, we found that pregnant women that defined their health status as poor and women without a previous COVID-19 diagnosis were more likely to get vaccinated against the COVID-19. In a similar way, Blakeway et al. (2022) found that pre-gestational diabetes mellitus is associated with COVID-19 vaccination uptake among pregnant women. Higher levels of fear about contracting COVID-19 increase the intention of individuals to accept a COVID-19 vaccine (Caserotti et al., 2021; Ward et al., 2020). For instance, intention of people to accept a COVID-19 vaccine is higher during the lockdown periods when the self-perceived COVID-19 vulnerability is higher (Brooks et al., 2020; Moccia et al., 2020). Moreover, past COVID-19 patients are less likely to be vaccinated, since they perceive a low risk and feel protected against the COVID-19 (Pacella-LaBarbara et al., 2021).

### Limitations

We should note a number of limitations in our study. Firstly, we conducted a cross-sectional study and therefore we are unable to establish a causal mechanism between psychosocial factors and COVID-19 vaccination uptake among pregnant women. Second, we relied on a convenience sample of pregnant women that cannot be considered representative of the population of pregnant women in Greece. For instance, the educational level of the participants in our study was high, while the participation rate of migrants was probably low since the questionnaire was only in Greek language. Furthermore, we used a valid questionnaire to measure psychosocial pattern of pregnant women but our data were based on self-reported measures which may introduce information bias due to tendency of participants to seek for social desirability. In addition, there are also other psychosocial factors that could affect pregnant women decision to receive a COVID-19 vaccine, e.g. anxiety, depression, quality of life, etc. Moreover, we did not measure some possible confounders, such as ethnicity, gestational week, at-risk pregnancy, vaccine type (mRNA or viral vector vaccine) that was offered to pregnant women, number of people in household, employment status (housewife or employed), and work location (working in person or working remotely).

### Conclusions

Our study is the first to assess psychosocial predictors of COVID-19 vaccines uptake among pregnant women with a valid instrument. This study is very timely due to the ongoing high COVID-19 case rates globally and the known increased risks of COVID-19 in pregnant women. An understanding of the psychosocial factors associated with increased COVID-19 vaccine uptake in pregnant women could be helpful for policy makers and healthcare professionals in their efforts to persuade women to get vaccinated against the COVID-19. Furthermore, we found that several socio-demographic characteristics of pregnant women affect their decision to receive a COVID-19 vaccine. Our findings emphasize the need for a more sensitive approach in the attempt to encourage vaccination uptake in pregnancy, especially in younger women, women with a good self-perceived health status, women with a previous COVID-19 diagnosis, and women who are not vaccinated against the seasonal influenza. Thus, public educational campaigns to encourage COVID-19 vaccination in the population of pregnant women should be targeted taking into account different concerns, needs, and attitudes. Mass vaccination of pregnant women is paramount to reduce COVID-19 vaccine hesitancy in pregnancy, halt spread of SARS-CoV-2 and reduce adverse outcomes in mothers and newborns.

## Data Availability

All data produced in the present study are available upon reasonable request to the authors

